# Remote monitoring of progression in early Parkinson’s disease: reliability and validity of the Roche PD Mobile Application v2

**DOI:** 10.1101/2021.10.07.21264414

**Authors:** Florian Lipsmeier, Kirsten I. Taylor, Ronald B. Postuma, Ekaterina Volkova-Volkmar, Timothy Kilchenmann, Brit Mollenhauer, Atieh Bamdadian, Werner L. Popp, Wei-Yi Cheng, Yan Ping Zhang, Detlef Wolf, Jens Schjodt-Eriksen, Anne Boulay, Hanno Svoboda, Wagner Zago, Gennaro Pagano, Michael Lindemann

## Abstract

Digital health technologies (DHTs) enable remote and therefore frequent measurement of motor signs, potentially providing reliable and valid estimates of motor sign severity and progression in Parkinson’s disease (PD). The Roche PD Mobile Application v1 was revised to v2 to include more measures of bradykinesia, and bradyphrenia and speech tests, to optimize suitability for early-stage PD. It was studied in 316 early-stage PD participants who performed daily active tests at home then carried a smartphone and wore a smartwatch throughout the day for passive monitoring (study NCT03100149). Adherence was excellent (96.29%). All pre-specified sensor features exhibited good-to-excellent test-retest reliability (median intraclass correlation coefficient = 0.9), and correlated with corresponding Movement Disorder Society - Unified Parkinson’s Disease Rating Scale items (rho: 0.12–0.71). These findings demonstrate the preliminary reliability and validity of remote at-home quantification of motor sign severity with the Roche PD Mobile Application v2 in individuals with early PD.

## Introduction

Clinical and drug development research in Parkinson’s disease (PD) critically requires the precise quantification of clinical disease severity and its progression over time. This is a challenge in PD because of the fluctuating and slowly progressive nature of the disease. Moreover, the development of disease-modifying therapies focus on the earliest possible point in the course of the disease, when the least amount of neurodegeneration has taken place, and when disease severity and progression may be even more subtle^1^. Digital health technologies (DHTs)^2^, such as smartphones and smartwatches, may aid in overcoming these challenges, since they enable remote and therefore frequent measurement of motor signs to provide potentially more robust – i.e. reliable and valid – quantification of disease severity and its changes over time^3-5^. Moreover, the inertial measurement unit sensors (e.g. accelerometers, gyroscopes) are highly sensitive to minute changes (even in consumer technologies) and therefore may detect motor changes not evident upon routine clinical examination. Here, we describe a novel DHT, the Roche PD Mobile Application v2, which was designed to measure motor manifestations in early PD, and demonstrate its test-retest reliability and validity in a group of de novo diagnosed individuals with PD.

Clinical trials of potential disease-modifying therapies for PD are especially promising among individuals who have been recently diagnosed, when less neurodegeneration has occurred. Clearly, to detect potential treatment benefit, sensitive measures of changes in motor signs in the earliest stages of the disease are required. In the Parkinson’s Progression Markers Initiative (PPMI) cohort^6^, Rasch Measurement Theory (RMT) analyses of the Movement Disorder Society - Unified Parkinson’s Disease Rating Scale (MDS-UPDRS)^7^ Part II and III scores at screening, 12-month and 24-month visits (n=384) revealed an apparent staged order of motor sign progression from unilateral bradykinesia and rigidity (first upper then lower extremities) to midline functions to bilateral bradykinesia and rigidity and finally general movement problems^8^. These findings confirm the centrality of bradykinesia in early PD^9^ and suggest that bradykinesia is a critical motor progression marker in early PD^1^. However, the quantification of bradykinesia and other motor signs in early PD with rating scales such as the MDS-UPDRS remains a challenge: MDS-UPDRS Part II scores change little in early PD^10^, i.e. less than the established minimal clinically meaningful difference^11^, and RMT analyses of MDS-UPDRS Part III item scores revealed multiple measurement irregularities in scores during the first 2 years of PPMI^8,12^. These findings highlight the urgent need for alternative methods of motor sign quantification in the earliest stages of PD.

Many smartphone- and smartwatch-based DHTs have been developed to estimate bradykinesia and other motor and non-motor signs of PD^5,13-19^. Finger tapping is one of the most commonly used DHT measures of bradykinesia^19^. When sensor-based finger-tapping data are aggregated over 2-week periods, test-retest reliabilities increase ^5^ and correlate with MDS-UPDRS Part III clinician ratings of finger-tapping performance in patients with early PD^5^. Additional bradykinesia tests implemented on smartphones include measures of hand turning and leg agility (by holding the phone on the thigh and lifting and stomping the foot), for example as implemented on CloudUPDRS^14,15^. Smartwatches offer additional means to estimate bradykinesia during daily life, for example by estimating the time taken to move a utensil from a plate to the mouth while eating.^13^ Most DHT solutions for PD such as CloudUPDRS^14,15^, HopkinsPD^16^, mPower^17^ and the Roche PD Mobile Application v1^20^ test not only bradykinesia, but also tremor, gait, and balance, thereby providing a profile of motor impairments for estimating and tracking PD motor severity. DHTs may additionally benefit from measures of cognition such as information processing speed (e.g., electronic Symbol Digit Modalities Test [eSDMT]) and speech, which are also affected in the earliest stages of PD^18^.

The present report describes the reliability and validity of the Roche PD Mobile Application v2, a revision of V1 to include multiple novel measures of bradykinesia and cognition (eSDMT and speech tests), and to optimize existing tasks for the detection of PD motor signs. Three hundred and sixteen individuals with early-stage PD (<2 years; Hoehn and Yahr stages I or II) who are participating in the Phase II PASADENA study (NCT03100149) were provided with a smartphone and smartwatch with the Roche PD Mobile Application v2 preinstalled, and requested to perform active tests daily on the smartphone (4 or 5 out of 10 tests each day, information processing speed once per fortnight), and to carry the smartphone and wear a smartwatch throughout the day to collect passive monitoring data. Pre-specified sensor features were calculated for each active test and for passive monitoring, and aggregated over the first two 2-week periods of the study. Adherence, test-retest reliability, and clinical validity (relationship to baseline clinical scales, known-groups validity) were quantified. These metrics represent the grounds for judging the potential utility of the Roche PD Mobile Application v2 to quantify and track progression of disease severity in early PD.

## Results

### Adherence

On average, daily remote active testing took a median of 5.3 (interquartile range [IQR] = 1.7) minutes on days without the SDMT, and 7.32 (median; IQR = 1.58) minutes on days with SDMT. Average adherence was high with 96.29% (median per participant) of all possible active tests performed during the first 4 weeks of the study. Participants contributed a median of 8.6 hours/day of smartphone and a median of 12.79 hours/day of smartwatch passive monitoring data.

### Reliability of sensor features

Reliability results are reported in Table **2**. All 17 pre-specified sensor features demonstrated good-to-excellent^21^ test-retest reliability between the first two 2-week study periods (ICCs ≥0.75) (Table **2**).^22^ The median sensor feature ICC was 0.9 (range, 0.75–0.95).

**Table 1.** Baseline characteristics of participants

|  | **Participants**<br>**(N=316)** |
| --- | --- |
| **Age, mean (SD), years** | 59.9 (9.10) |
| **Male, n (%)** | 213 (67%) |
| **MDS-UPDRS Part IA, mean (SD)** | 1.16 (1.62) |
| **MDS-UPDRS Part IB, mean (SD)** | 3.45 (2.78) |
| **MDS-UPDRS Part II, mean (SD)** | 5.33 (4.04) |
| **MDS-UPDRS Part III, mean (SD)** | 21.47 (9.00) |
| **MDS-UPDRS Part VI, mean (SD)** | NA |
| **MDS-UPDRS Total Score, mean (SD)** | 31.41 (12.78) |
| **PIGD score, mean (SD)** | 1.04 (0.90) |
| **Bradykinesia score, mean (SD)** | 10.50 (5.71) |
| **Rigidity score, mean (SD)** | 4.58 (2.95) |
| **Tremor score, mean (SD)** | 5.56 (4.01) |
| **Speech score, mean (SD)** | 2.72 (2.19) |
| **Axial symptoms, mean (SD)** | 2.04 (1.71) |
| **Hoehn and Yahr stage, %** |  |
| Stage I | 32 |
| Stage II | 67 |
| Stage III | 1 |

MDS-UPDRS, Movement Disorder Society - Unified Parkinson's Disease Rating Scale;
NA, not applicable; PIGD, Postural Instability/Gait Disorders; SD, standard deviation.

**Table 2.** Reliability and clinical validity of pre-specified Roche PD Mobile Application v2 active test and passive monitoring sensor features. “M” = most affected side; “L” = least affected side

| Digital test | Sensor feature | MDS-UPDRS item | Test-retest reliability | Spearman's correlation coefficient |  | MDS-UPDRS item score 0 vs 1 |  |  |  |  | Hoehn and Yahr Stage 1 vs. 2 |  |  |  | Feature differences in expected direction |
| --- | --- | --- | --- | --- | --- | --- | --- | --- | --- | --- | --- | --- | --- | --- | --- |
| | | | ICC (95% CI) | $r_s$ | *P* value | n | U stat | *P* value | N 0/N 1 | Percent difference on group medians | U stat | *P* value | N 1/N 2 | Percent difference on group medians | |
| Draw A Shape | Spiral celerity (L) | 3.5 Hand movement (L) | 0.91<br>(0.88–0.93) | –0.15 | 0.0092 | 300 | 7061 | 0.0043 | 184/95 | –14.06 | 6681 | 0.0047 | 74/226 | –9.5 | Yes |
|  | Spiral celerity (M) | 3.5 Hand movement (M) | 0.92<br>(0.89–0.94) | –0.12 | 0.0334 | 300 | 3091 | 0.0981 | 49/144 | –14.29 | 6463 | 0.0017 | 74/226 | –15.2 | Yes |
| Dexterity | Tapping variability (L) | 3.4 Finger tapping (L) | 0.84<br>(0.79–0.88) | 0.32 | < 0.0001 | 300 | 6786 | 0.0006 | 152/116 | 27.33 | 6444 | 0.0015 | 74/226 | 27.16 | Yes |
|  | Tapping variability (M) | 3.4 Finger tapping (M) | 0.88<br>(0.84–0.91) | 0.37 | < 0.0001 | 300 | 1261 | 0.0006 | 30/135 | 40.64 | 6691 | 0.005 | 74/226 | 17.95 | Yes |
| Hand Turning | Median Hand Turning speed (L) | 3.6 Pronation-supination (L) | 0.92<br>(0.89–0.94) | –0.18 | 0.0015 | 300 | 7930 | 0.0361 | 192/95 | –7.85 | 7268 | 0.0458 | 74/226 | –7.18 | Yes |
|  | Median Hand Turning speed (M) | 3.6 Pronation-supination (M) | 0.95<br>(0.93–0.97) | –0.46 | < 0.0001 | 300 | 1912 | 0.0008 | 43/131 | –12.37 | 7216 | 0.0385 | 74/226 | –9.33 | Yes |
| Speech | MFCC2 variability | 3.1 Speech | 0.77<br>(0.69–0.83) | –0.29 | < 0.0001 | 295 | 5692 | < 0.0001 | 126/138 | –13.68 | 6395 | 0.0412 | 71/209 | –4.47 | Yes |
| Phonation | Voice jitter | 3.1 Speech | 0.88<br>(0.85–0.91) | 0.18 | 0.0017 | 297 | 7381 | 0.0001 | 135/146 | 21.65 | 7248 | 0.073 | 73/224 | 12.87 | Yes |
| Postural Tremor | Log median squared energy (L) | 3.15 Postural tremor (L) | 0.83<br>(0.77–0.87) | 0.23 | < 0.0001 | 300 | 3240 | 0.0002 | 259/39 | 8.26 | 7111 | 0.0343 | 73/227 | 1.2 | Yes |
|  | Log median squared energy (M) | 3.1 Postural tremor (M) | 0.92<br>(0.90–0.94) | 0.5 | < 0.0001 | 300 | 4779 | < 0.0001 | 150/116 | 12.45 | 8108 | 0.3918 | 73/227 | 1.28 | Yes |
| Rest Tremor | Log median squared energy (L) | 3.17 Rest tremor amplitude (L) | 0.91<br>(0.88–0.93) | 0.34 | < 0.0001 | 300 | 967 | < 0.0001 | 274/19 | 23.73 | 6614 | 0.0048 | 73/227 | 6.12 | Yes |
|  | Log median squared energy (M) | 3.17 Rest tremor amplitude (M) | 0.94<br>(0.91–0.95) | 0.71 | < 0.0001 | 300 | 1565 | < 0.0001 | 124/71 | 19.27 | 7896 | 0.2731 | 73/227 | 3.59 | Yes |
| Balance | Log sway jerk | 3.12 Postural stability | 0.82<br>(0.76–0.87) | 0.14 | 0.0138 | 299 | 890 | 0.0071 | 287/11 | 10.6 | 7988 | 0.3870 | 72/227 | 1.29 | Yes |
| U-turn | Median turn speed | 3.14 Body bradykinesia | 0.91 | –0.22 | 0.0001 | 299 | 3113 | 0.0539 | 44/168 | –5.39 | 6322 | 0.0019 | 72/227 | –7.83 | Yes |
|  |  |  | (0.88–<br>0.93) |  |  |  |  |  |  |  |  |  |  |  |  |
| eSDMT | Number of<br>correct<br>responses | 1.1 Cognitive<br>impairment | 0.75<br>(0.66–<br>0.81) | –0.18 | 0.0014 | 303 | 3787.5 | 0.0012 | 248/43 | –10 | 6538 | 0.0008 | 76/227 | –7.14 | Yes |
| Passive<br>monitoring<br>of gait | Median<br>turn speed | 3.14 Body<br>bradykinesia | 0.82<br>(0.77–<br>0.89) | –0.22 | 0.0001 | 285 | 3130 | 0.154 | 43/162 | –3.3 | 6270 | 0.0138 | 71/214 | –4.65 | Yes |
| Passive<br>monitoring<br>of gesture | Median<br>gesture<br>power | 3.6 Pronation-supination, left<br>hand | 0.85<br>(0.80–<br>0.89) | –0.43 | < 0.0001 | 269 | 3108 | < 0.0001 | 106/89 | –35.73 | 5221 | 0.0242 | 60/209 | –30.64 | Yes |

CI, confidence interval; eSDMT, electronic Symbol Digit Modalities Test; ICC, intraclass correlation coefficient; L, less affected side;
M, more affected side; MDS-UPDRS, Movement Disorder Society - Unified Parkinson's Disease Rating Scale; MFCC2, Mel Frequency
Cepstral Coefficient 2.

### Clinical validity of sensor features

Clinical validity was assessed via Spearman’s correlations between the sensor features and corresponding MDS-UPDRS subscale and item scores (Fig. **1**). Correlations with MDS-UPDRS item scores revealed that all sensor features correlated with their corresponding clinical items (Table **2** and Figs. **3**–**6**). The numerically strongest relationships were observed with bradykinesia sensor features (i.e. Hand Turning and Finger Tapping) as well as postural and rest tremor sensor features, particularly for the more affected side, while the numerically weakest correlations were found with the Balance (comparator: MDS-UPDRS item 3.12 Postural stability) and Draw A Shape (comparator: MDS-UPDRS item 3.5 Hand movement) tests.

**Fig. 1.**
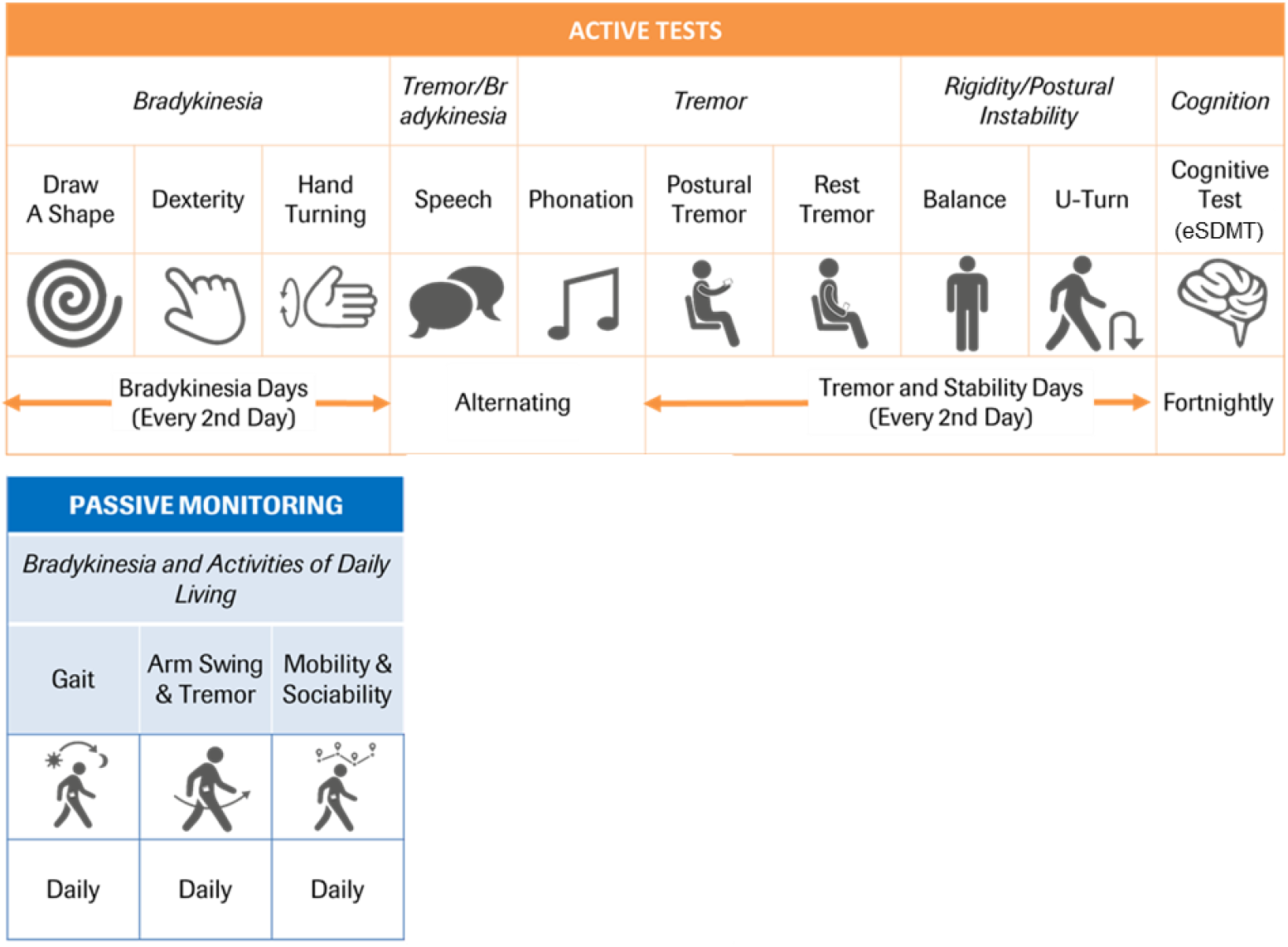
Roche PD Mobile Application v2 active tests and passive monitoring and schedule of assessments eSDMT, electronic Symbol Digit Modalities Test; PD, Parkinson’s disease.

**Fig. 2.**
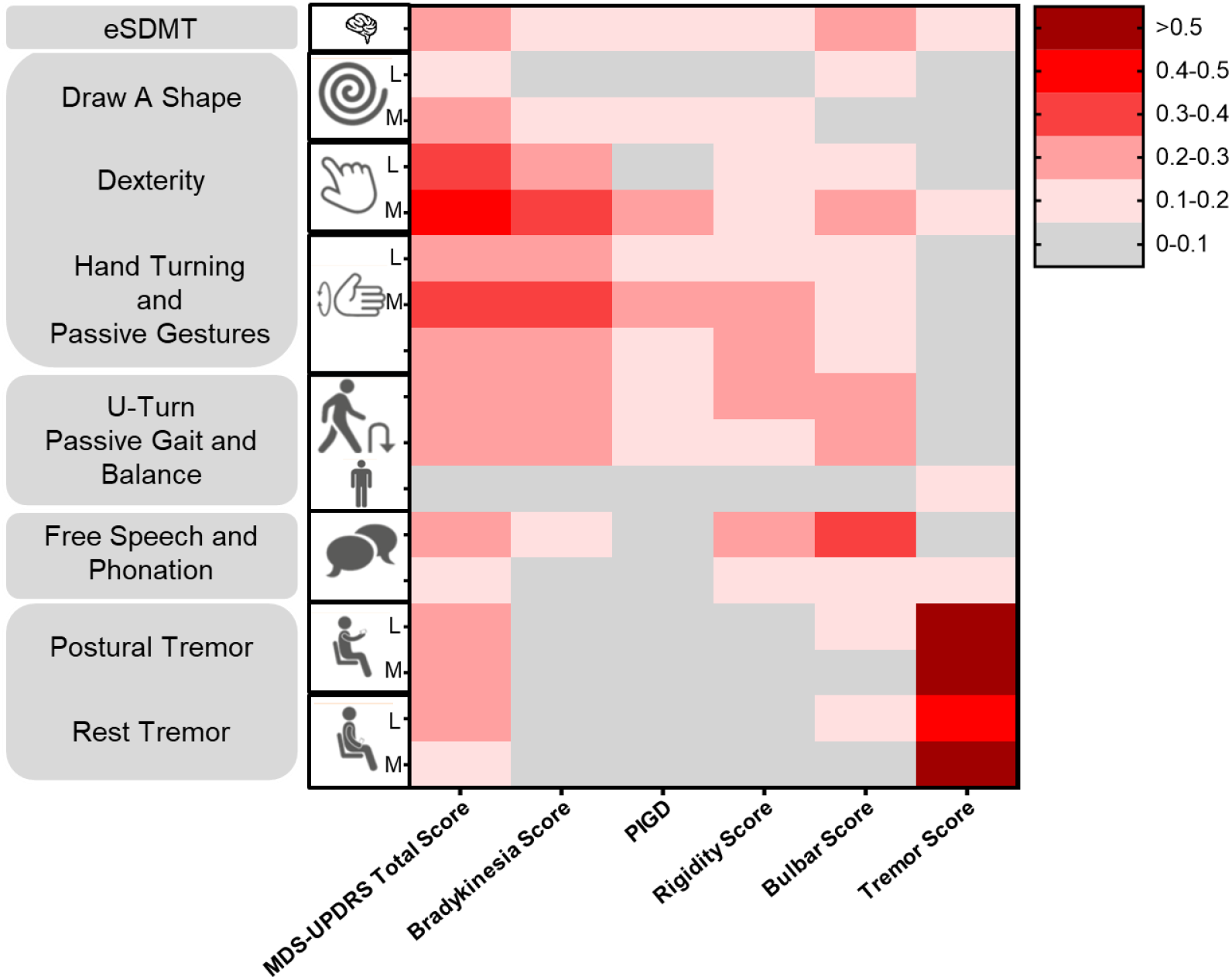
Absolute Spearman’s correlations between baseline MDS-UPDRS Total and Sub-scores and Roche PD Mobile Application v2 active test and passive monitoring sensor features eSDMT, electronic Symbol Digit Modalities Test; MDS-UPDRS, Movement Disorder Society - Unified Parkinson’s Disease Rating Scale; PD, Parkinson’s disease; PIGD, Postural Instability/Gait Disorders.

**Fig. 3.**
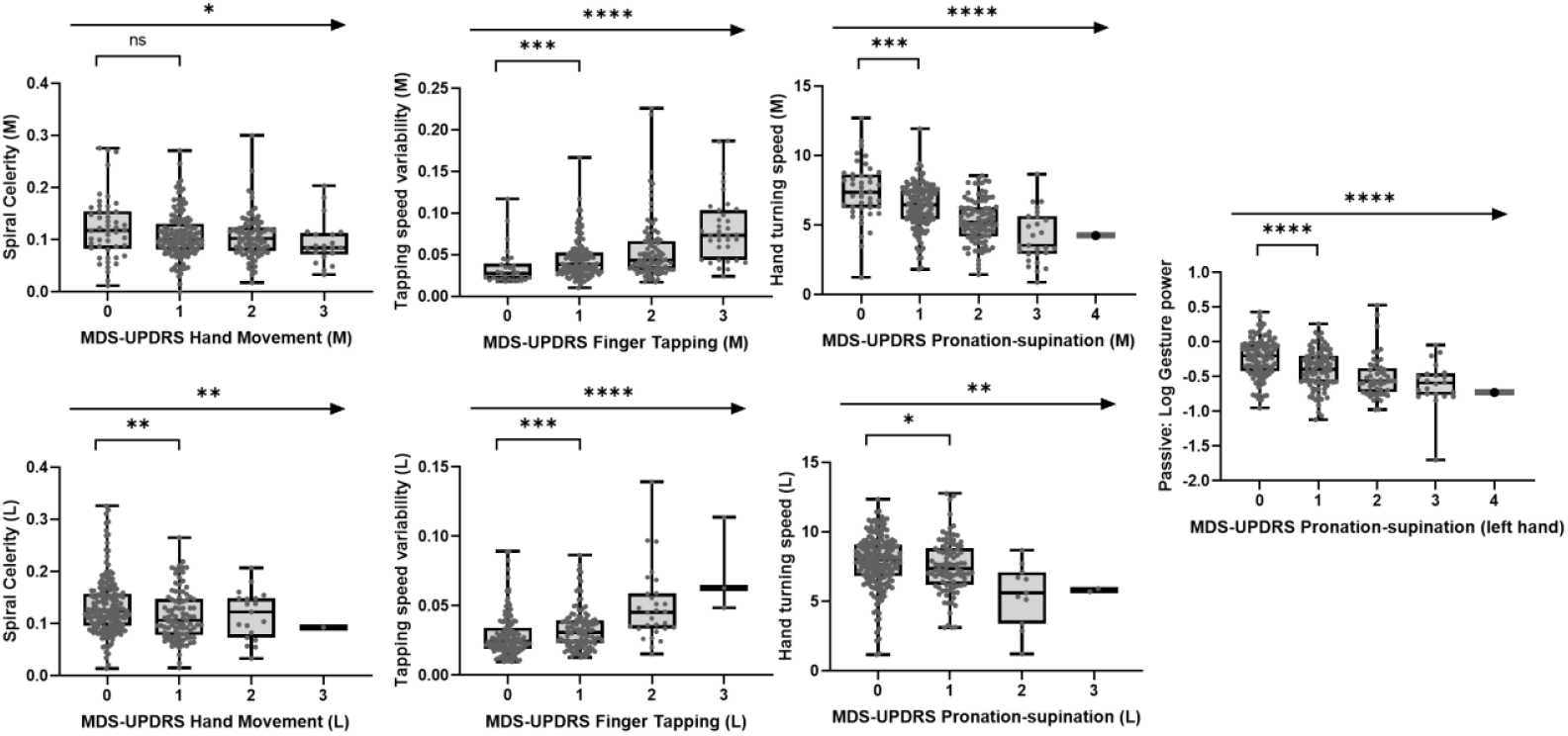
Association of sensor features from upper limb bradykinesia tests with corresponding clinical MDS-UPDRS measures at baseline (ns = *P* > 0.05; * = *P* ≤ 0.05; **= *P* ≤ 0.01; *** = *P* ≤ 0.001; ****= *P* ≤ 0.0001) L, less affected side; M, more affected side; MDS-UPDRS, Movement Disorder Society - Unified Parkinson’s Disease Rating Scale.

**Fig. 4.**
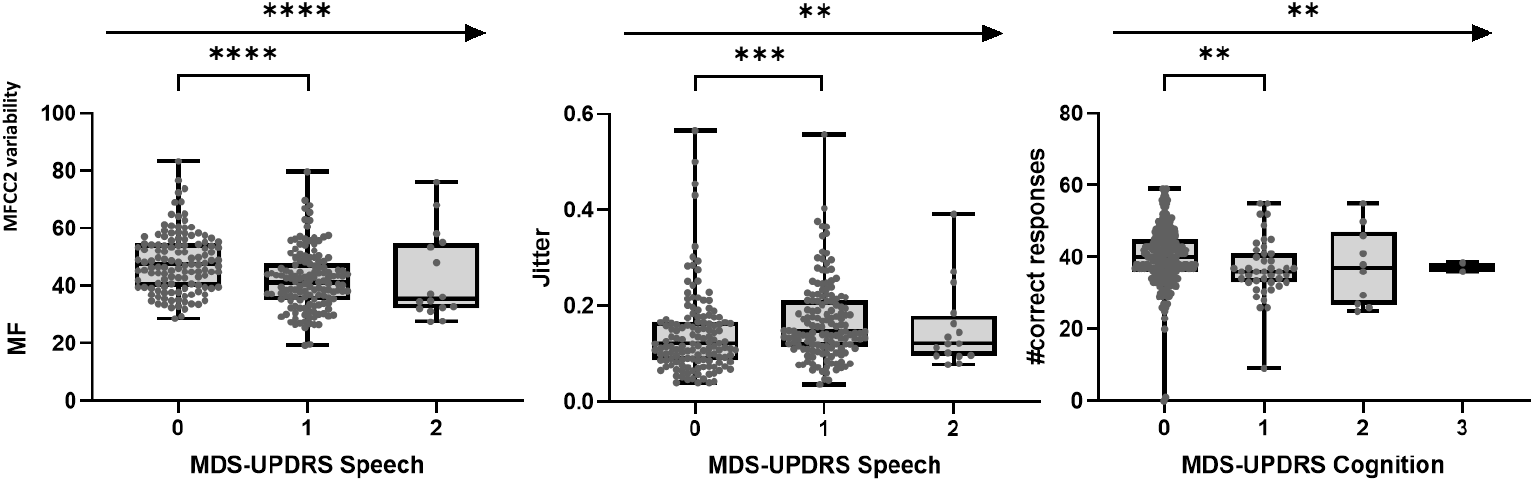
Association of sensor features from Phonation/Speech and eSDMT tests with corresponding clinical MDS-UPDRS measures at baseline (ns = *P* > 0.05; * = *P* ≤ 0.05; **= *P* ≤ 0.01; *** = *P* ≤ 0.001; ****= *P* ≤ 0.0001) eSDMT, electronic Symbol Digit Modalities test; MDS-UPDRS, Movement Disorder Society - Unified Parkinson’s Disease Rating Scale; MFCC2, Mel Frequency Cepstral Coefficient 2.

**Fig. 5.**
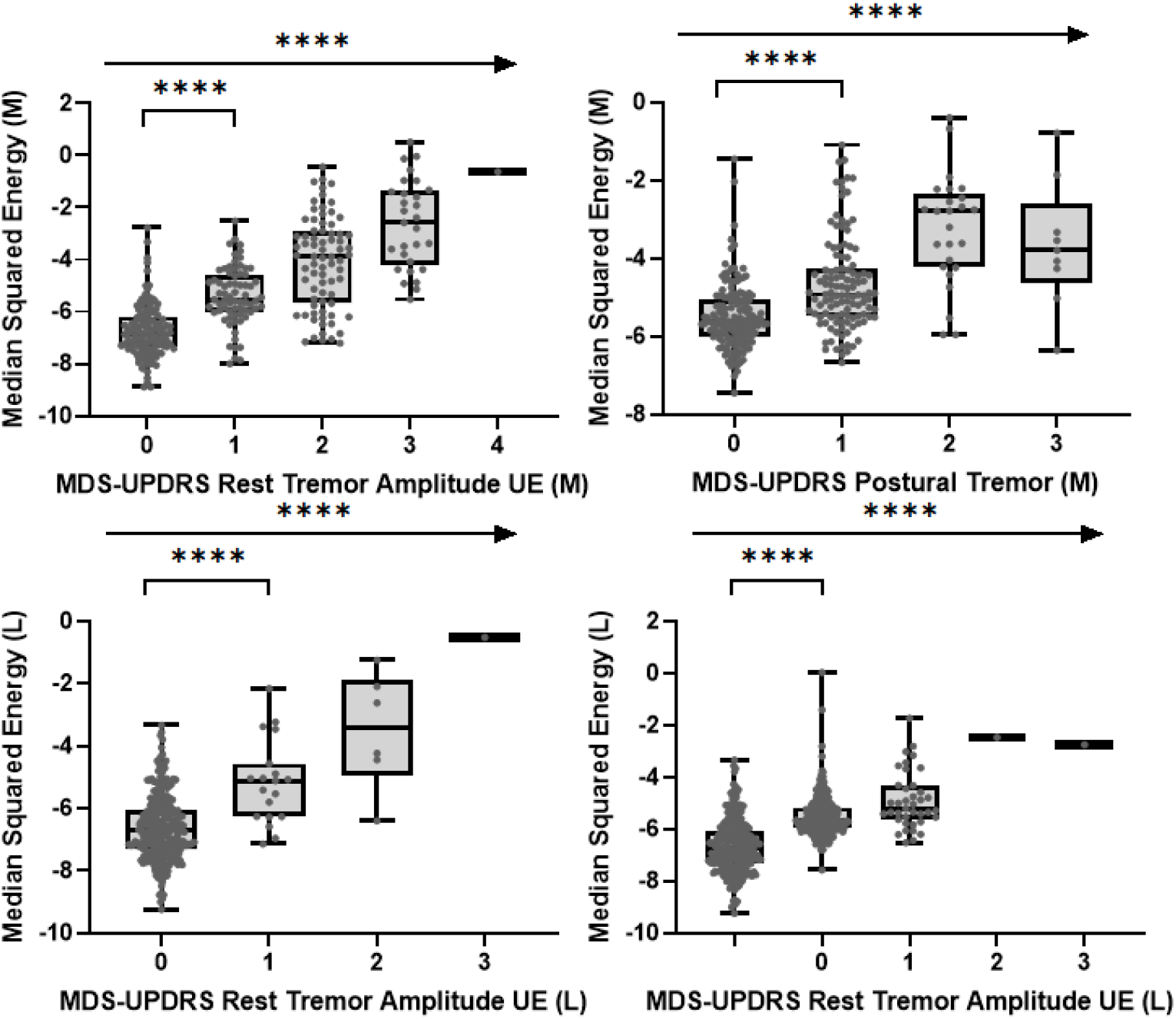
Association of sensor features from upper limb tremor tests with corresponding clinical MDS-UPDRS measures at baseline (ns = *P* > 0.05; * = *P* ≤ 0.05; **= *P* ≤ 0.01; *** = *P* ≤ 0.001; ****= *P* ≤ 0.0001) L, less affected side; M, more affected side; UE, Upper Extremity; MDS-UPDRS, Movement Disorder Society - Unified Parkinson’s Disease Rating Scale.

**Fig. 6.**
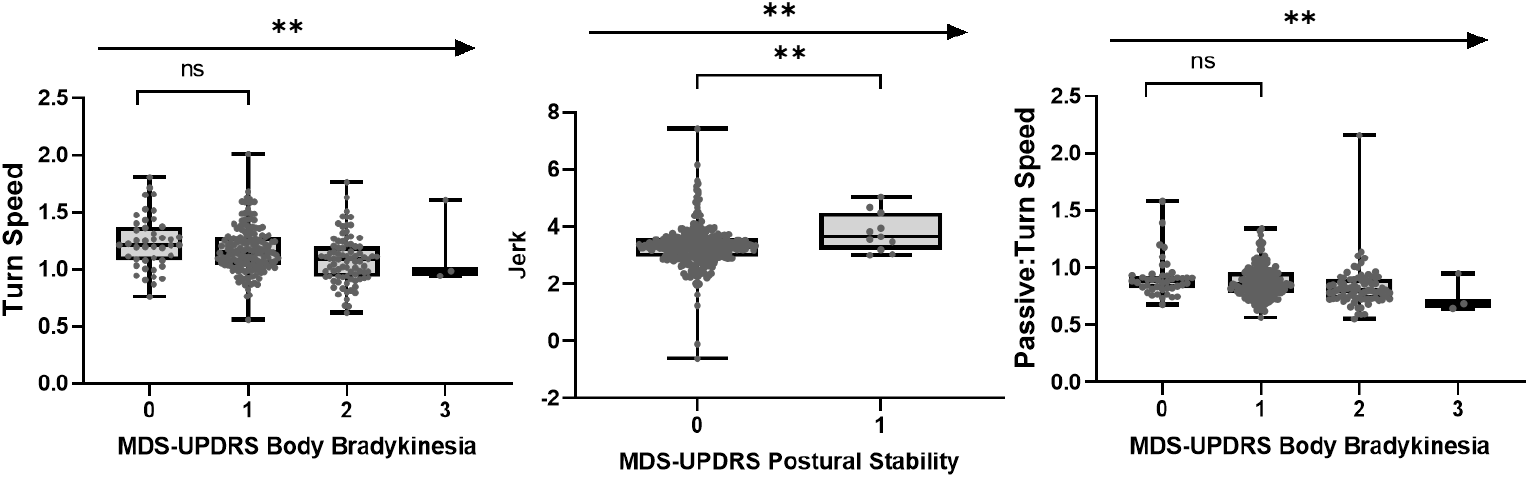
Association of sensor features from U-turn, Balance tests and Passive monitoring of gait with corresponding clinical MDS-UPDRS measures at baseline (ns = *P* > 0.05; * = *P* ≤ 0.05; **= *P* ≤ 0.01; *** = *P* ≤ 0.001; ****= *P* ≤ 0.0001) MDS-UPDRS, Movement Disorder Society - Unified Parkinson’s Disease Rating Scale.

Cross-correlations between sensor features and MDS-UPDRS subscale scores demonstrated convergent and divergent validity of bradykinesia and tremor sensor features, with the strongest relationships between bradykinesia sensor features and MDS-UPRS subscores, and between tremor sensor features and MDS-UPDRS subscores, compared with all other subscores (Fig. **2**). The Postural Instability/Gait Disorders (PIGD) subscale score showed the strongest correlations with the U-turn and the Hand Turning test (most affected side). Rigidity subscale scores correlated weakly with all sensor features overall, showing a weak-to-moderate relationship with Hand Turning in the most affected side and U-turn speed. The novel ‘bulbar score’ (defined in the methods section below) correlated most strongly with the Speech Test sensor feature. Overall, MDS-UPDRS total scores were most strongly related to bradykinesia sensor features, as expected for this early-stage population^1^.

### Sensor feature sensitivity to early disease manifestations and Hoehn and Yahr stage

A total of 15/17 sensor features discriminated participants with scores of 0 versus 1 on associated MDS-UPDRS items (Table **2** and Figs. **3**–**6**). Only sensor features from the Draw a Shape and U-turn tests showed borderline non-significant results. Participants in Hoehn and Yahr Stage I versus II were differentiated by 13 sensor features, i.e. all but Phonation test, Postural and Rest Tremor (most affected side) and the Balance test sensor features (Table **2**).

### Sensor feature sensitivity to side differences

Sensor features values from all lateralized tests demonstrated significant differences between the most and least affected sides (Table **3**). Moreover, sensor features and MDS-UPDRS scores measuring the same motor sign on the less affected (or more affected) side were more strongly correlated than sensor features and MDS-UPDRS scores measuring the same motor sign on different sides of the body (Fig. **7**).

**Table 3.** Differences in sensor feature values from lateralized active tests between less affected and more affected sides. (green color indicates that the difference is in the expected direction)

| Active test | U stat | *P* value | Percent difference |
| --- | --- | --- | --- |
| **Rest Tremor** | 29543 | < 0.0001 | 12.97 |
| **Postural Tremor** | 34696 | < 0.0001 | 6.89 |
| **Hand Turning** | 23025 | < 0.0001 | -22.7 |
| **Dexterity** | 25818 | < 0.0001 | 50.78 |
| **Draw A Shape** | 35909 | < 0.0001 | -13.45 |

**Fig. 7.**
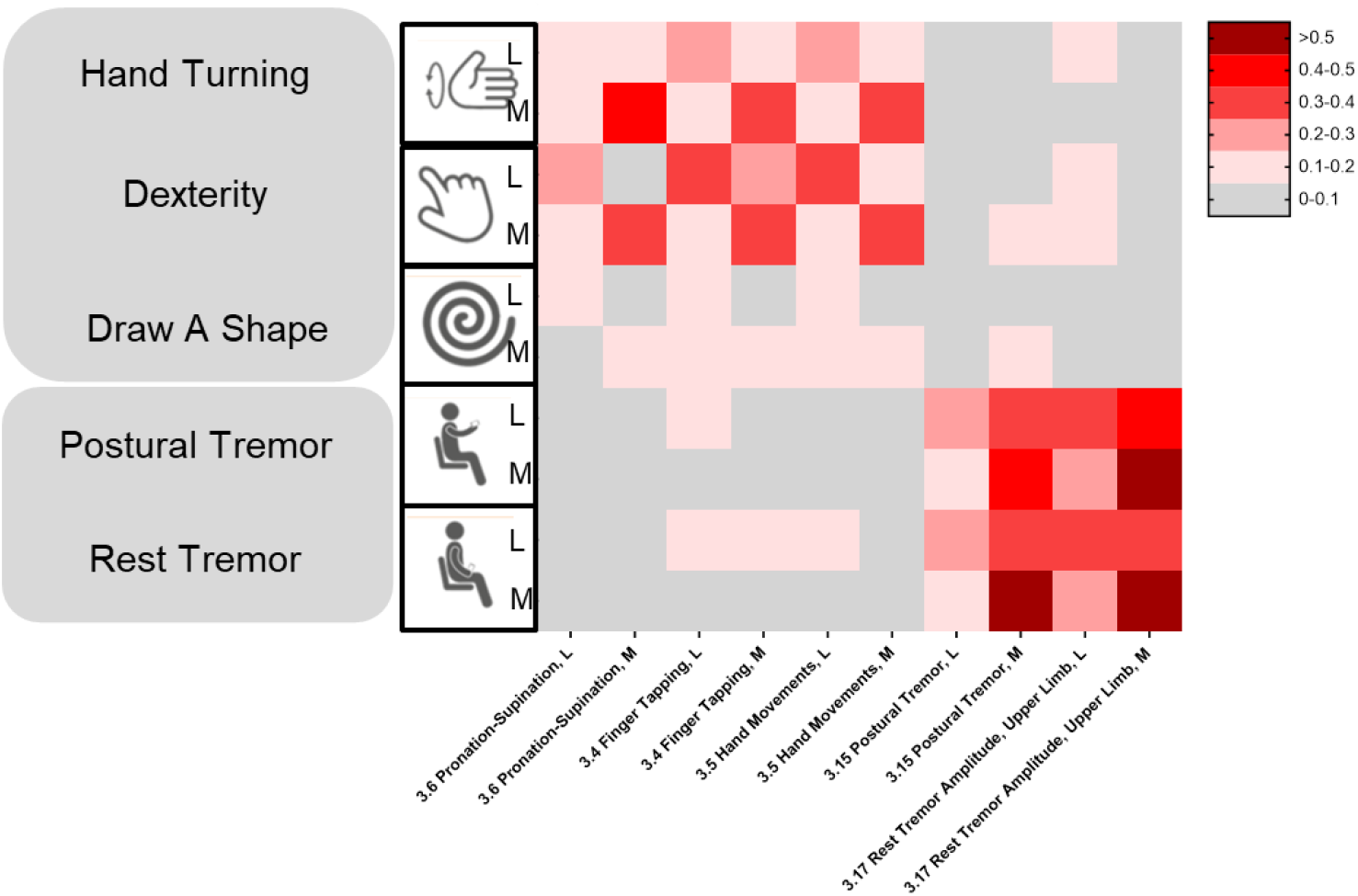
Correlations between active test sensor features from the more and less affected sides (M and L, respectively) with MDS-UPDRS Part III item scores evaluating M and L L, less affected side; M, more affected side; MDS-UPDRS, Movement Disorder Society - Unified Parkinson’s Disease Rating Scale.

## Discussion

The Roche PD Mobile Application v1 was designed to measure the core motor signs of PD, and was recently revised to v2 to primarily include two new active tests of bradykinesia (Hand Turning, Draw A Shape), as well as a test of psychomotor slowing (eSDMT) and a speech test. In addition, the original gait task was revised to a U-turn test, and a smartwatch was incorporated into the remote passive monitoring procedure. Preliminary test-retest reliability scores for the pre-specified sensor features from all active tests except Speech and eSDMT, and for both passive monitoring measures, were in the ‘excellent’ range^21^. Preliminary clinical validity was established via correlations with corresponding MDS-UPDRS item scores. We note that these findings are reassuring considering the continuous (sensor feature) versus ordinal (MDS-UPDRS) nature of the two datasets, and the lack of conceptually comparable MDS-UPDRS items for some active test features (e.g. Draw A Shape). Cross-correlations between sensor features and MDS-UPDRS subscale scores supported the convergent and divergent validity of bradykinesia and tremor sensor features. Most active test sensor features demonstrated sensitivity for subtle manifestations, discriminating individuals who received MDS-UPDRS item scores of 0 from those with item scores of 1. Measures of upper limb bradykinesia demonstrated known-groups validity, differentiating individuals in Hoehn and Yahr Stage I versus II. All lateralized sensor features discriminated least versus the most affected side of the body. Taken together, these results indicate that the Roche PD Mobile Application v2 may prove suitable for quantifying motor disease severity and tracking disease progression in the earlier stages of PD.

The Roche PD Mobile Application v2 contains three active tests designed to measure upper limb bradykinesia: Dexterity (finger tapping), Hand Turning (pronation/supination), and Draw A Shape. Pre-specified sensor features from all three tests correlated with their corresponding MDS-UPDRS upper limb bradykinesia item scores, and showed convergent and divergent validity in cross-correlations with MDS-UPDRS Part III subscale scores, correlating numerically most strongly with bradykinesia compared with all other subscale scores. These findings indicate that the Roche PD Mobile Application v2 bradykinesia tests indeed reflect the neurological concept of upper limb bradykinesia. Finger tapping and pronation/supination tasks are well-established assessments of upper limb bradykinesia as evidenced by their inclusion in both the UPDRS^23^ and MDS-UPDRS^7^. Over the last decade, different digitized variants of finger tapping and pronation/supination tests have been developed^19^. Despite methodological differences, studies of these DHT tasks generally showed good correspondence between finger tapping sensor features and respective clinical ratings, as well as the ability to differentiate healthy controls from individuals with early PD, and individuals with early PD from individuals with later-stage PD^16,20,24-27^, in line with the present findings. While the literature on digitized pronation/supination assessments is less rich than for finger tapping, available results also consistently demonstrate correlations with related clinical scores and the ability to differentiate healthy participants from individuals with PD^14,28-31^. Spiral drawing is traditionally used in behavioral neurology to assess fine motor impairment including bradykinesia and tremor^32-35^. DHT versions of spiral drawing demonstrated that time to completion correlated with clinician ratings of bradykinesia severity, and differentiated PD cases from controls^34^. The majority of previous DHT spiral drawing tasks used pens/digital pens to draw on regular paper or tablets, a more challenging motor task compared with the present finger drawing on smaller smartphone touch screens. In the present study, celerity, i.e. accuracy/time to complete spiral shape tracing on the smartphone screen, was pre-specified to additionally consider the accuracy of directed fine motor movements in the unsupervised at-home setting. Spiral celerity correlated with MDS-UPDRS bradykinesia measures, and the strength of these correlations was numerically smaller compared with Finger Tapping and Hand Turning. This may be due to the relative difficulty of the latter two tasks compared with spiral drawing, which may have challenged individuals more, thereby revealing greater impairment. We note that additional sensor features (e.g. variability in drawing speed, hesitation), analyzed either individually or combined within and across shapes, are expected to provide additional meaningful information, as has been shown for PD and multiple sclerosis^36,37^.

Passive monitoring with smartwatches provides a unique opportunity to explore slowing of upper limb movements during daily life. Here, sensor data segments during gesture movements were identified from the circa 90% non-walking periods in the passive monitoring sensor data stream, using the squared magnitude of the accelerometer sensor movement as the sensor feature. This same feature has been related to decreased expressivity in patients with schizophrenia with negative symptoms^38^. Here, gesture power was specifically related to the MDS-UPDRS bradykinesia subscore and item scores, as well as the rigidity subscore, and is in line with a slowing of hand movement in daily non-gait-related activities such as gesturing when speaking, eating, etc. These findings are consistent with previous research with wrist-worn wearables, which traditionally focused on arm swing during gait^39-41^, as well as multi-sensor systems used to measure the impact of bradykinesia on activities of daily living^13,42^. Thus, passively monitored motor behavior in daily life may facilitate our understanding of the effect and burden of PD on individuals’ daily lives.

The eSDMT^43^ is commonly applied to measure psychomotor slowing, or bradyphrenia, one of the earliest cognitive signs in PD, appearing up to 5 years prior to a PD dementia diagnosis^18^. However, as the test requires multiple cognitive functions, it is not surprising that it is sensitive to many forms of neurologic impairment.^44^ Indeed, while SDMT performance is reduced in PD^45^, impairments are exacerbated in individuals with PD with concomitant vascular^46^ and amyloid^47^ imaging findings. A standard SDMT outcome measure,- number of correct responses in 90 seconds, was pre-specified for the present analyses of the eSDMT, and showed ‘good’^21^ test-retest reliability (ICC = 0.75). However, it correlated only weakly (rho = –0.18) with the MDS-UPDRS item 1.1. assessing global cognitive impairment. This finding is surprising given the catch-all nature of both the eSDMT and MDS-UPDRS item 1.1., but may be accounted for by the fact that cognitive impairments were excluded during the screening process in the PASADENA study, leading to a truncation of range in both scores (see Supplementary Fig. **1**). We note that we attempted to minimize the effect of bradykinesia on eSDMT scores by requiring a simple tap response on a number pad displayed at the bottom half of the smartphone screen. Nevertheless, to mitigate the risk of this confound, eSDMT performance could be controlled by a non-cognitively demanding motor test using a similar response format.

Voice and speech impairments in PD are varied and generally summarized under the term dysarthria, and include resonatory, articulatory, phonatory, prosodic and respiratory components^48^. This symptomatology and its relevance to patients’ daily lives motivated the inclusion of a Sustained Phonation task in the suite of active tests, and the development of the novel Speech test. Voice jitter was pre-selected as a proxy of disordered vocal fold function for the sustained phonation test. In line with previous research^49^, increased voice jitter correlated weakly with MDS-UPDRS 3.1. (Speech) scores, and differentiated individuals with slight speech disturbances (MDS-UPDRS 3.1. score of 1) from those with no perceivable speech impairment at the site visit (MDS-UPDRS 3.1. score of 0). In the Speech active test, monotonicity (i.e., MFCC2 fundamental frequency variability) was selected as the sensor feature of prosodic deficits based on previous research demonstrating that this feature differentiated individuals with PD from healthy controls^48^. In the present study, MFCC2 variability correlated with MDS-UPDRS 3.1. (Speech) scores, and differentiated participants with MDS-UPDRS 3.1. scores of 0 and 1. The bulbar MDS-UPDRS Part III composite item score was designed to gauge the severity of motor impairments in body parts involved in speech production. Despite a truncation of range in this score (average <3/20 points), MFCC2 variability correlated with the bulbar score, indicating that this feature may estimate the severity of motor impairments in the speech apparatus. Future research will investigate further richly multi-faceted aspects of speech function to better understand motor and cognitive behavior in PD.

The Roche PD Mobile Application v2 aims to assess the broad array of motor signs in PD and related movement disorders. Thus, besides bradykinesia, speech, voice, and psychomotor slowing, tremor (rest, postural), turning during gait, and balance were also assessed. The rest and postural tremor active test features corresponded most strongly to the respective MDS-UPDRS concepts of tremor, as demonstrated by the highest correlation overall with any MDS-UPDRS item and subscale scores. This is consistent with similar DHT reports^5,24,50^. The novel U-turn test (which instructed individuals, if safe to do so, to walk several paces and make a U-turn at least five times) and the identification of turning while walking throughout the day in passive monitoring sensor data, were motivated by findings that turning is particularly impaired in PD^5,51,52^. For example, a 360 degree walking turn and instrumented timed-up-and-go test showed strong reliability and discriminated controls from PD participants^53,54^. Similarly, sensor-based measures of turn speed in daily life differentiated PD individuals from controls^55^. In the present study, turn speed measured in both the active test and passive setting correlated with MDS-UPDRS 3.14. body bradykinesia item scores, but was not specifically related to MDS-UPDRS PIGD relative to other subscores. While neither measure of turn speed differentiated between less and more affected individuals on MDS-UPDRS body bradykinesia scores of 0 versus 1, both differentiated between individuals in Hoehn and Yahr Stage I versus II. Although participants were not instructed to ‘turn as fast as possible’ to ensure a safe conduct of the active test, the U-turn test showed numerically higher correlations with body bradykinesia compared with passive turning speed, in line with similar profile of performance (active testing) versus capacity (passive monitoring) scores previously demonstrated for gait speed^56^. In the balance active test, the jerk sensor feature correlated with the MDS-UPDRS 3.12. postural stability item score, similar to previous reports^5,57^, and differentiated individuals with MDS-UPDRS item 3.12 scores of 0 versus 1, but failed to differentiate individuals in Hoehn and Yahr Stage I versus II. We speculate that this negative finding may reflect the low levels of gait and postural instability impairments in the present cohort (mean PIGD=1).

A composite summary score of individual features across diverse assessments is expected to provide a more robust measure of global PD severity and progression, especially given the heterogeneous nature of PD. Several DHT solutions besides the Roche PD Mobile Application v2 administer different motor active tests, and some additionally collect passive monitoring data^5,15,16^. Supplementary Table **1** provides a high-level comparison of these DHT solutions. All solutions contain active tests for tremor and tapping, but vary with respect to the inclusion of other upper limb, postural stability/gait, cognition, and voice/speech tests, and whether passively monitored motor data are collected. The power of combining different features across the tests in these DHTs has been shown via machine learning models that predict MDS-UPDRS total scores (Roche PD Mobile Application v1)^58^ or lead to a new score based on differentiation of ON and OFF L-dopa states^59^, and distinguished between healthy controls, idiopathic Rapid Eye Movement and PD^14,60^. A machine learning approach was also used to combine different HopkinsPD baseline sensor features to predict clinically significant events (e.g. falls, functional impairment) at the 18-month follow-up^61^. In contrast to data-driven approaches to composite score development, a clinical outcomes assessment approach could be applied whereby information from individuals with PD inform the selection of sensor features such that they optimally reflect what matters most to patients^62^.

Several facets of the present study limit the generalizability of the findings. Firstly, all individual’s disease duration was <2 years, and individuals were in Hoehn and Yahr Stages I or II. Thus, the applicability of the present findings to later-stage or prodromal PD is unknown. The reduced range of disease severities also appeared to limit the ranges of some DHT and clinical measures, which consequently limited the possibility to detect relationships between the two (Supplementary Fig. **1**). Second, since Roche PD Mobile Application v2 data are not yet available from neurologically normal individuals, sensor feature cut-off values differentiating normal from impaired motor behavior could not yet be calculated. It should be also noted that comparisons between DHT measures and clinical measures such as the MDS-UPDRS can also be affected by limitations in the clinical measures; if an active test is not adequately reflected by a clinical measure, the ability to detect meaningful correlations is reduced. Finally, only two continuous 2-week periods of DHT data were analyzed; thus, the long-term adherence to the remote monitoring procedure and ability of sensor features to detect changes over time remain to be established. Towards this end, it is critical to quantify and report test-retest reliabilities of sensor feature scores towards assessing a sensor feature’s potential to detect changes over time^63^ and any deviation from normal progression as a function of e.g. pharmacological interventions.

The Roche PD Mobile Application v2 was designed to measure the severity of early PD core motor signs and to provide information complementary to established clinical outcome measures. This remote monitoring approach enables high-frequency (i.e. daily) assessments with low average daily burden. The frequent measurement coupled with the high sensitivity of smartphone/smartwatch sensors may increase signal-to-noise of digital outcome measures for clinical research and provide novel insights into patients’ functioning in daily life.

## Methods

### Participants

Baseline Roche PD Mobile Application v2 data from 316 dopaminergic-treatment-naïve individuals recently diagnosed with dopamine transporter imaging with single-photon emission computed tomography-confirmed PD (Hoehn and Yahr Stages I–II, diagnosis ≤2 years) were analyzed (see Table **1** for demographic and clinical characteristics). All individuals were enrolled in an ongoing randomized, double-blind, placebo-controlled, Phase II clinical trial (PASADENA Part 1; NCT03100149) of prasinezumab (RO7046015/PRX002), an anti-α-synuclein monoclonal antibody (see Pagano G et al, 2021)^64^.

All of the >60 PASADENA sites received approval from their institutional review boards or ethics committees, and written informed consent was provided by all participants. The study is being conducted in accordance with the Declaration of Helsinki and the International Conference on Harmonization Guidelines for Good Clinical Practice.

### Roche PD Mobile Application v2

The Roche PD Mobile Application v2 consists of an application installed on a provisioned smartphone and smartwatch (see Fig. **1**). The PD Mobile Application prompted participants to perform the active tests described below. All unilateral tests were performed twice, once with each side of the body.

1. Draw A Shape:^37^ participants were instructed to trace six different shapes (two diagonal lines [once drawn up, once drawn downwards], and a square, circle, figure-8, and spiral) on the smartphone screen with their index finger, as quickly and as accurately as possible (timeout: 30 seconds);
2. Dexterity: participants were instructed to alternately tap two touchscreen buttons with their index finger as quickly and regularly as possible (20 seconds per hand);
3. Hand Turning: holding the smartphone in the outstretched hand while seated, participants were instructed to rotate the hand as quickly and fully as possible such that the phone faced up and down (10 seconds per hand);
4. Speech: participants were provided with three open-ended questions, one after the other, and instructed to read each out loud and answer each question out loud (20 seconds per question);
5. Phonation: participants were instructed to make a single, continuous “aaaah” sound for as long as possible with one breath and in a steady pitch and volume while the phone was held at the ear (timeout: 30 seconds);
6. Postural tremor: participants were instructed to sit with their eyes closed, and to hold the smartphone in an outstretched hand while counting down out loud from a pre-specified number that differed for each test administration (15 seconds per hand);
7. Rest tremor: participants were instructed to sit with their eyes closed and to hold the phone in the palm of their hand, with their forearm resting on their thigh, and to count down out loud from a pre-specified number that differed for each test administration (15 seconds per hand);

8.Balance: while standing with the smartphone in a running belt around their waist, participants were instructed to stand still with their arms at their side (30 seconds);

9.U-turn: participants were instructed to place the smartphone in a running belt, and to walk between two points at least four steps apart at normal speed, completing at least five turns in 60 seconds;

10.eSDMT^43^: participants were instructed to match a sequence of displayed symbols to the respective numbers using a displayed coding key as quickly and as accurately as possible (90 seconds).

The Roche PD Mobile Application v2 additionally administered questionnaires, which are not the focus of the present report. For passive monitoring, participants were instructed to carry their smartphone (e.g. in their trouser pocket or in the pouch of a provided running belt) and wear their smartwatch as they conducted the daily active tests and their normal daily activities. No active tests were administered directly via the smartwatch.

### Procedure

Participants were provided with an Android smartphone (Galaxy S7, Samsung, Seoul, South Korea) and smartwatch (Moto G 360 2nd Gen Sport; Motorola, Chicago, USA) during a screening visit at the latest 7 days prior to the baseline clinical visit, and trained on the use of the devices and the Roche PD Mobile Application v2. Participants were instructed to open the application on the provisioned smartphone every morning. Active tests were scheduled automatically such that half of the motor tests were presented on alternating days, and the eSDMT every 2 weeks (Fig. **1**), with a total expected testing time including transitions and test-start countdowns between tests of 5–10 minutes (including eSDMT).

Baseline clinical assessments included the MDS-UPDRS^7^, from which subscale scores were generated (i.e. PIGD, bradykinesia, rigidity, tremor)^26^. Additionally, a ‘bulbar score’ was defined as the composite sum of MDS-UPDRS items 2.1 Speech; 2.2 Saliva and Drooling; 2.3 Chewing and Swallowing; 3.1 Speech; and 3.2 Facial expression.

### Sensor data processing

The raw sensor data from the smartphone and smartwatch were extracted and converted into pre-defined ‘sensor features’, one per active test performed and side of body (if applicable) and one for passive monitoring. Features were selected based on previous literature and their relevance to PD (Supplementary Table **2**).

Data underwent quality control (QC) checks to ensure that the tests had been performed properly. Towards this end, QC metrics were generated. For example, one QC metric quantified the amount of energy from the accelerometer during the Hand Turning test to estimate whether the smartphone was lying still (e.g. on a table) or moving during the test. 0.3% (n=179/56,786) of digital active test data not meeting the pre-specified QC thresholds and were therefore excluded from the analyses.

### Statistical analyses

Sensor features from passive monitoring and each active test performed were summarized (median) over 2-week intervals starting at the baseline visit (Weeks 1 and 2) and in the 2-week period thereafter, provided that ≥3 data points were available during each 2-week testing interval (Supplementary Table **3**). For convergent validity, the averaged (median) sensor data collected during the first two study weeks were compared with clinical data collected at the baseline visit (Day 1) using Spearman’s correlations. Adherence and test-retest metrics were calculated for aggregated sensor features for the first two 2-week study periods. Adherence was defined as the number of fully completed active testing sessions relative to the number of all possible active testing sessions. For passive monitoring, the average number of hours per day participants carried the smartphone with them and wore the smartwatch was calculated. Sensor feature test-retest reliabilities were quantified with the ICC between averaged values of the first and second 2-week periods. To investigate the sensitivity of sensor features to subtle or very early symptoms, sensor features from participants receiving MDS-UPDRS item scores of 0 versus 1 were compared using Mann-Whitney U tests. For known-groups validity, sensor features were compared between participants in Hoehn and Yahr Stage I versus II, and by comparing sensor feature values from less and more affected sides, both using Mann-Whitney U tests.

## Supporting information

Ethics checklist

## Data Availability

Qualified researchers may request access to individual patient-level data through the clinical trial data request platform (https://vivli.org/). Further details on Roche's criteria for eligible trials are available at https://vivli.org/members/ourmembers/. For further details on Roche's Global Policy on the Sharing of Clinical Information and how to request access to related clinical trial documents, see https://www.roche.com/research_and_development/who_we_are_how_we_work/clinical_trials/our_commitment_to_data_sharing.htm.

## Acknowledgements

We gratefully acknowledge the participants of the PASADENA study and their families, as well as the PASADENA Investigators. We also acknowledge the Roche PD Mobile application v2 development team, especially Hans van Wesenbeeck, Juraj Korcek, Monnika Broennimann for their contributions in the development and deployment of the solution. The authors thank Sarah Child and Kristina Rodriguez, PhD, of MediTech Media for providing editorial support and Megan Speakman of MediTech Media for medical editing assistance.

## Author information

### Contributions

FL, KIT, RBP, TK, BM, and ML designed the study. TK, BM, AB, WLP, JSE, and JB were involved in data collection. FL, KIT, EVV, WLP, WC, YZ, DW, and ML analysed the data. FL, KIT, RBP, EVV, HS, WZ, GP and ML were involved in data interpretation. FL, KIT, and ML were involved in drafting the work. All authors revised and gave input on the article. All authors approved the final content and are accountable for all aspects of the publication.

### Ethics declarations

Participants were identified for potential recruitment using site-specific recruitment plans prior to consenting to take part in this study. Recruitment materials for participants had received Institutional Review Board or Ethics Committee approval prior to use. The following Institutional Review Boards ruled on ethics of the PASADENA study: Ethikkommission der Medizinischen Universität Innnsbruck, Innsbruck, Austria; Comité de Protection des Personnes (CPP) Ouest IV, Nantes, France; Ethikkommission der Universität Leipzig and Geschäftsstelle der Ethikkommission an der medizinischen Fakultät der Universität Leipzig, Leipzig, Germany; Ethikkommission der Fakultät für Medizin der Technischen Universität München, München, Germany; Ethikkommission der Universität Ulm (Oberer Eselsberg), Ulm, Germany; Landesamt für Gesundheit und Soziales Berlin and Geschäftsstelle der Ethik-Kommission des Landes Berlin, Berlin, Germany; Ethikkommission des FB Medizin der Philipps-Universität Marburg, Marburg, Germany; Ethikkommission an der Medizinischen Fakultät der Eberhard-Karls-Universität und am Universitätsklinikum Tübingen, Tübingen, Germany; Ethikkommission an der Med. Fakultät der HHU Düsseldorf, Düsseldorf, Germany; Ethikkommission der LÄK Hessen, Frankfurt, Germany; CEIm Hospital Universitari Vall d’Hebron, Barcelona, Spain; Copernicus Group Independent Review Board, Puyallup, Washington, USA; Western Institutional Review Board, Puyallup, Washington, USA; The University of Kansas Medical Center Human Research Protection Program, Kansas City, Kansas, USA; Oregon Health & Science University Independent Review Board, Portland, Oregon, USA; Northwestern University Institutional Review Board, Chicago, Illinois, USA; Spectrum Health Human Research Protection Program, Grand Rapids, Michigan, USA; The University of Vermont Committees on Human Subjects, Burlington, Vermont, USA; Beth Israel Deaconess Medical Center Committee on Clinical Investigations, New Procedures and New Forms of Therapy, Boston, Massachusetts, USA; Vanderbilt Human Research Protection Program Health, Boston, Massachusetts, USA; Vanderbilt Human Research Protection Program Health, Nashville, Tennessee, USA; University of Maryland, Baltimore Institutional Review Board, Baltimore, Maryland, USA; University of Southern California Institutional Review Board, Los Angeles, California, USA; Columbia University Medical Center Institutional Review Board, New York, New York, USA; University of Southern California San Francisco Institutional Review Board, San Francisco, California, USA; University of Pennsylvania Institutional Review Board, Philadelphia, Philadelphia, USA; HCA - HealthOne Institutional Review Board, Denver, Colorado, USA. All Institutional Review Boards gave ethical approval of the study.

