## Supplementary materials for "Remote monitoring of progression in early Parkinson’s disease: reliability and validity of the Roche PD Mobile Application v2"

**Supplementary Table 1. Smartphone- and smartwatch-based active test suites reported which were tested in more than 30 participants with PD**

| Domain | Test | mPower | HopkinsPD | CloudUPDRS | Roche v1* | Roche v2** |
|---|---|---|---|---|---|---|
| Tremor upper limb | Rest Tremor | | x | x | x | x |
| | Postural Tremor | | x | x | x | x |
| | Kinetic Tremor | | | x | | |
| Tremor lower limb | Rest Tremor | | | x | | |
| Upper limb bradykinesia | Hand turning | | | x | | x |
| | Finger tapping[22,66] | x | x | x | x | x |
| | Draw A Shape[37] | | | | | x |
| Gait/postural instability | Balance | | x | | x | x |
| | U-turn | | | | | x |
| | Gait | x | x | x | x | x |
| | Leg agility | | | x | | |
| Voice/Speech | Sustained phonation | x | x | | x | x |
| | Speech | | | | | x |
| Cognition | eSDMT[18] | | | | | x |
| | Memory | x | | | | |
| | Reaction time | | x | | | |
| Passive monitoring phone | | | x | | x | x |

| | | | | | | |
|---|---|---|---|---|---|---|
| Passive monitoring watch | | | | | | x |

* Roche PD Mobile Application v1.

** Roche PD Mobile Application v2.

eSDMT, electronic Symbol Digit Modalities test; PD, Parkinson’s disease; UPDRS, Unified Parkinson’s Disease Rating Scale.

**Supplementary Fig. 1. Distributions of MDS-UPDRS parts and subscores at baseline clinical visit**

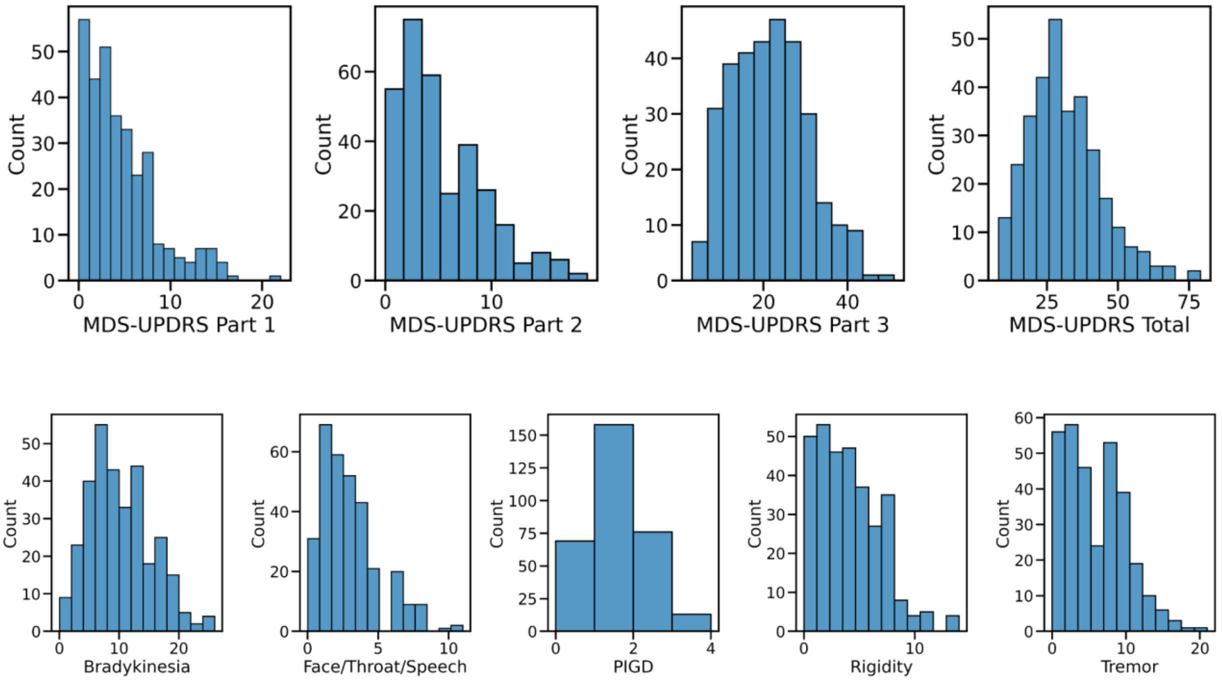


MDS-UPDRS, Movement Disorder Society - Unified Parkinson's Disease Rating Scale; PIGD, Postural Instability/Gait Disorders.

**Supplementary Table 2. Pre-specified sensor feature for each active test and for passive monitoring**

| DHT assessment | Sensor feature | Description |
|---|---|---|
| Draw A Shape | Spiral celerity | Accuracy of drawing the spiral relative to drawing duration. Hypothesis: higher celerity indicates less impairment |
| Dexterity | Tapping variability | Standard deviation of tapping speed. Hypothesis: higher variability indicates more impairment |
| Hand Turning | Median Hand Turning speed | Median of turn speed over all hand rotations from the gyroscope. Hypothesis: higher hand turning turn speed indicates less impairment |
| Speech | MFCC2 variability | Standard deviation of average MFCC2 for words. Hypothesis: higher variability corresponds to less monotonicity which indicates less impairment |
| Phonation | Voice jitter | Mean absolute difference between the period of adjacent pitch cycles, normalized by the mean pitch period, multiplied by 100. Hypothesis: higher jitter indicates more impairment |
| Rest and postural tremor | Log median squared energy | Median of the acceleration magnitude. Hypothesis: higher energy corresponds to higher tremor amplitude indicating more impairment |
| Balance | Log sway jerk | Jerk of the acceleration magnitude. Hypothesis: more jerk indicates more impairment |
| U-turn | Median turn speed | Median over turn speed per turn from the gyroscope. Hypothesis: higher turn speed indicates less impairment |
| SDMT | Number of correct responses | Number of correctly matched symbol digits. Hypothesis: more correct responses indicates less impairment |
| Passive monitoring (smartphone, gait) | Median turn speed in passive monitoring | Median of turn speed over all detected turns during 1 day of recording. Hypothesis: higher turn speed indicates less impairment |
| Passive monitoring (smartwatch, gestures) | Median gesture power | Median of integrated squared acceleration magnitude over all identified gestures during 1 day of recording. Hypothesis: higher gesture power indicates less impairment |

MFCC2, Mel Frequency Cepstral Coefficient 2; SDMT, Symbol Digit Modalities Test.
